# Community-Based Participatory Approaches in Implementing HIV Stigma Reduction Interventions in Healthcare Settings: A Qualitative Study from Indonesia

**DOI:** 10.64898/2025.12.08.25341857

**Authors:** Mona S Mahmud, Rafdzah A Zaki, Rumana A Saifi, Theresia P. Kusumoputri, Devika Devika, Dean Retno, Frederick L Altice, Adeeba Kamarulzaman

## Abstract

**Background:** This study investigated the effectiveness of a community-based participatory approach (CBPA) in implementing the *“Intervensi Penghapusan Stigma dan Diskriminasi”* (IPSD) intervention to reduce HIV-related stigma among healthcare workers in Indonesian primary healthcare settings. The IPSD training reached 3,162 participants across 42 primary health centres and 56 referral hospitals. The intervention engages people with HIV (PWH) and key populations (KP) as co-developers and co-implementers. Despite recognised challenges in addressing HIV-related stigma in healthcare settings, evidence on CBPA effectiveness in Indonesia remains limited.

**Methods:** To assess the effectiveness of CBPA in executing the IPSD intervention, 13 semi-structured, in-depth interviews and 7 focus group discussions were conducted with 40 participants, including policymakers, healthcare professionals, and community expert trainers (CETs). Thematic analysis was employed, with interview questions grounded in the Consolidated Framework for Implementation Research (CFIR) and Proctor’s implementation outcomes framework. Data analysis was facilitated using NVivo 14, with codes systematically categorised to identify concepts aligned with CFIR constructs.

**Results:** Findings reveal that CBPA fostered a more inclusive healthcare environment, demonstrated by improved provider attitudes toward people with HIV and reduced discrimination. CBPA enabled collaboration between healthcare providers, PWH, and KPs through CETs as external change agents. This study shows CBPA’s potential in reducing HIV-related stigma and calls for broader adoption to achieve equitable healthcare, particularly in settings with HIV stigma.

**Conclusion:** Findings underscore the transformative potential of community participation stigma reduction interventions in public healthcare settings. By promoting structured contact and shared learning between stigmatised and non-stigmatised groups, the CBPA strengthened provider attitudes, enhanced implementation quality, and advanced equity within Indonesia’s healthcare system.

## Background

Indonesia faces considerable challenges in meeting the UNAIDS 95-95-95 targets for HIV treatment. In 2022, it was estimated that 570,000 individuals were living with HIV (PWH) in Indonesia, with a prevalence rate of 0.4%(1). Current data indicates that 60% of PWH are aware of their status, 41% are receiving antiretroviral therapy (ART), and 38% have achieved viral suppression (1). Despite a reduction in HIV infections, there has been an increase in AIDS-related mortality, attributed to deficiencies in treatment coverage. The national HIV program emphasises the decentralisation of HIV services from hospitals to primary health centres (PHCs) to enhance community-based care (2). The limited adoption of ART adversely affects prevention efforts and heightens the risk of disease progression. Although treatment is administered in clinical settings, sustained care necessitates collaboration between healthcare providers and the community. Stigma within healthcare settings poses a critical threat to HIV control initiatives, undermines patient retention and adversely affects the quality of life of PWH (3–5).

## HIV Stigma in the Indonesian Healthcare Settings

Laws and policies have been established to promote non-discrimination toward PHIV, ensuring equality, protection of confidentiality, and informed consent when accessing services. However, the Global Fund report found significant and severe stigma against individuals perceived to belong to KP groups. The report acknowledged that stigma and discrimination in healthcare facilities are not uniform, with varying degrees of severity depending on the facility’s location (6).

Stigma and discrimination related to HIV within Indonesian healthcare settings are shaped by individual beliefs, peer influences, and patient characteristics(7). HIV-related stigma associated with HIV is linked to a limited understanding of its transmission and prevention (8–10), as well as factors such as gender, health discipline, socioeconomic status, and religious beliefs (10). The implementation of a supportive workplace culture characterised by diversity and inclusive policies can mitigate stigmatising attitudes (8, 11).

The PWH Stigma Index report, which surveyed 1,400 respondents, indicated that 19.5% of individuals living with HIV encountered stigma within HIV-specific services and 15.9% experienced it in non-HIV healthcare services. Instances of avoidance of physical contact were reported by 4.3% of respondents in HIV services and 5.6% in non-HIV services. The study further revealed that 16.8% of people who use drugs (PWUD), 16.7% of transgender people (TG), and 12.6% of female sex workers (FSW) avoided healthcare facilities because of anticipated stigmatisation. Although 86.9% of participants started ART immediately, 7.2% delayed initiation, and 11% felt pressured by healthcare workers. Additionally, 58.9% of key populations (KPs) postponed ART initiation due to concerns about disclosing their HIV status (12).

### IPSD Stigma Reduction Intervention

In 2018, Indonesia’s Ministry of Health (MoH) initiated the “Intervensi Penghapusan Stigma dan Diskriminasi” (IPSD) intervention to address HIV-related stigma among HCWs. This initiative was adapted from the “Health4ALL Health Workers’ Training Guide for the Provision of Quality, Stigma-free HIV Services for Key Populations”. The Health4All training guide was created under the FHI360-LINKAGES project to enhance the capacity of healthcare workers to deliver equitable, comprehensive, and stigma-free HIV services to key populations. The IPSD intervention was conducted in 42 primary health centres (PHCs) through in-person training and in 56 referral hospitals via online training, reaching a total of 3,162 participants.

The guide uses participatory adult learning methodologies and emphasises self-reflection, empathy, and values clarification to improve clinical and interpersonal competencies. It promotes an inclusive and rights-based approach to care by encouraging providers to examine their attitudes and biases, better understand the lived experiences of KPs, and implement actionable changes in the delivery of services.

The Health4All training guide emphasizes the importance of community engagement in its implementation. Consequently, a community-based participatory approach (CBPA) was used to implement the IPSD, involving PWH and representatives of KPs as resource persons and community expert trainers (CETs).

### Community-based Participatory Approaches (CBPA)

CBPA interventions create supportive environments through multilevel strategies, leveraging community strengths to ensure that health initiatives are tailored and effective (13, 14). These approaches have been successful in improving health-seeking behaviours, access to services, and health literacy, particularly among socio-economically disadvantaged populations (15). The lack of meaningful community involvement has hindered the dissemination of many public health initiatives (16).

Research indicates that educational and structural interventions, such as providing antiretroviral therapy (ART) and implementing supportive government policies, effectively reduce HIV-related stigma (17). Participatory training interventions have shown promise in decreasing stigma in healthcare settings (18). Furthermore, positive interactions between healthcare professionals and patients enhance adherence to treatment (19). Although interventions have successfully reduced stigma in various health contexts, their widespread implementation and sustainability in low– and middle-income countries remain limited (Kemp et al., 2019). Current stigma reduction research predominantly focuses on individual-level interventions, with a notable lack of evidence supporting community engagement in national initiatives (3).

### Research Objectives

This study investigated CBPA’s effectiveness in planning and implementing the IPSD stigma reduction intervention. The research question was: “How did the community-based participatory approach strategy facilitate the planning and implementation of the stigma and discrimination reduction (IPSD) intervention?”. This broad question avoids limiting the scope of the enquiry (20). This study employed a deductive approach, which relies on pre-established theories to guide the design and expects the findings to corroborate valid theories (21).

## Methods

### Study Design

This study investigates the initial implementation outcomes identified by Proctor et al., specifically adoption, focusing on short-term outcomes assessed qualitatively through in-depth interviews (IDIs) and focus group discussions (FGDs). The development of semi-structured interview questions was informed by the Consolidated Framework for Implementation Research (CFIR) and Proctor’s Implementation Outcomes Framework. Data analysis was performed using NVivo 14, with codes categorised to identify the concepts corresponding to the CFIR constructs.

The interview guide consisted of ten primary questions and twenty-one sub-questions, delving into various aspects of implementation and focusing on the experiences of stakeholders, including PHIV and KP experts (Appendix 1). Health facilities and participants were selected through consultations with the MoH and PHIV network. From a total of 42, ten primary health centres (PHCs) were purposively selected for IDI and FGD, representing five provincial administrative cities in Greater Jakarta.

Interviews were conducted via Zoom from March to June 2023 in Bahasa Indonesia, each lasting 60-90 minutes. Participants were compensated with a Rp100,000 (USD 6.50) internet package. The recorded audio files were transcribed and translated into English before being imported into NVivo 14 for coding and analysis. The translated interview transcripts were coded separately in NVivo 14 by the principal investigator (PI), co-PI, and one research assistant (RA). The PI consolidated the coded files and reviewed them for consistency.

The analysis employed the CFIR as the theoretical model, using pre-selected constructs to code observed phenomena. The codes were categorised based on their similarities to identify recurring concepts. While primarily deductive, the analysis also inductively explored emerging themes. Thematic analysis identified patterns that were organised in Microsoft Excel, revealing themes aligned with the CFIR constructs.

Ethical considerations included respect for individuals sharing sensitive information, such as sexual orientation and HIV status. Recruitment through MoH and PHIV Networks emphasised voluntary participation. Confidentiality was maintained through secure Zoom sessions and restricted file-sharing. Following the CBPA, the PI collaborated with PHIV networks and KP representatives, including a community representative, for data collection. Consent forms outlining the study details were obtained before the interviews, emphasising voluntary participation (Appendix 2).

### Ethical Approval

This study was reviewed and approved by the Ethics Committee of the Institute of Research and Community Service, Atma Jaya Catholic University, Indonesia (approval number 001D/III/PPE). PM.10.05/01/2023. All participants were informed about the purpose, procedures, and voluntary nature of the study. Written informed consent was obtained from all participants prior to data collection. Confidentiality and anonymity were ensured throughout the research process in accordance with the ethical guidelines for human subjects’ research.

## Results

### Participant Characteristics

A total of forty (n=40) participants engaged in semi-structured IDIs and FGDs (Table 1). The cohort comprised policymakers, international partners, Heads of Primary Health Centres (PHCs), HCWs, and community expert trainers (CETs).

**Table 1:**
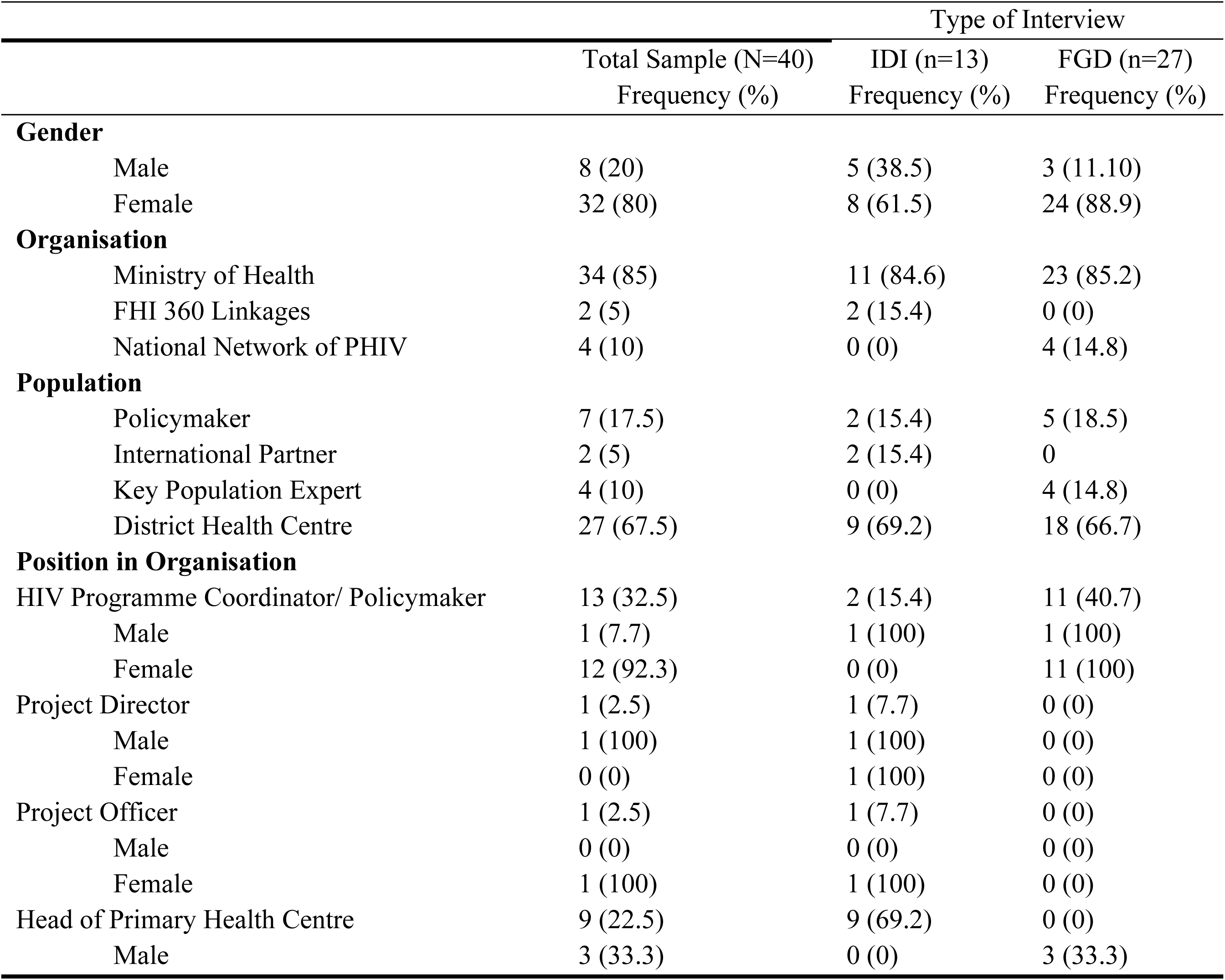
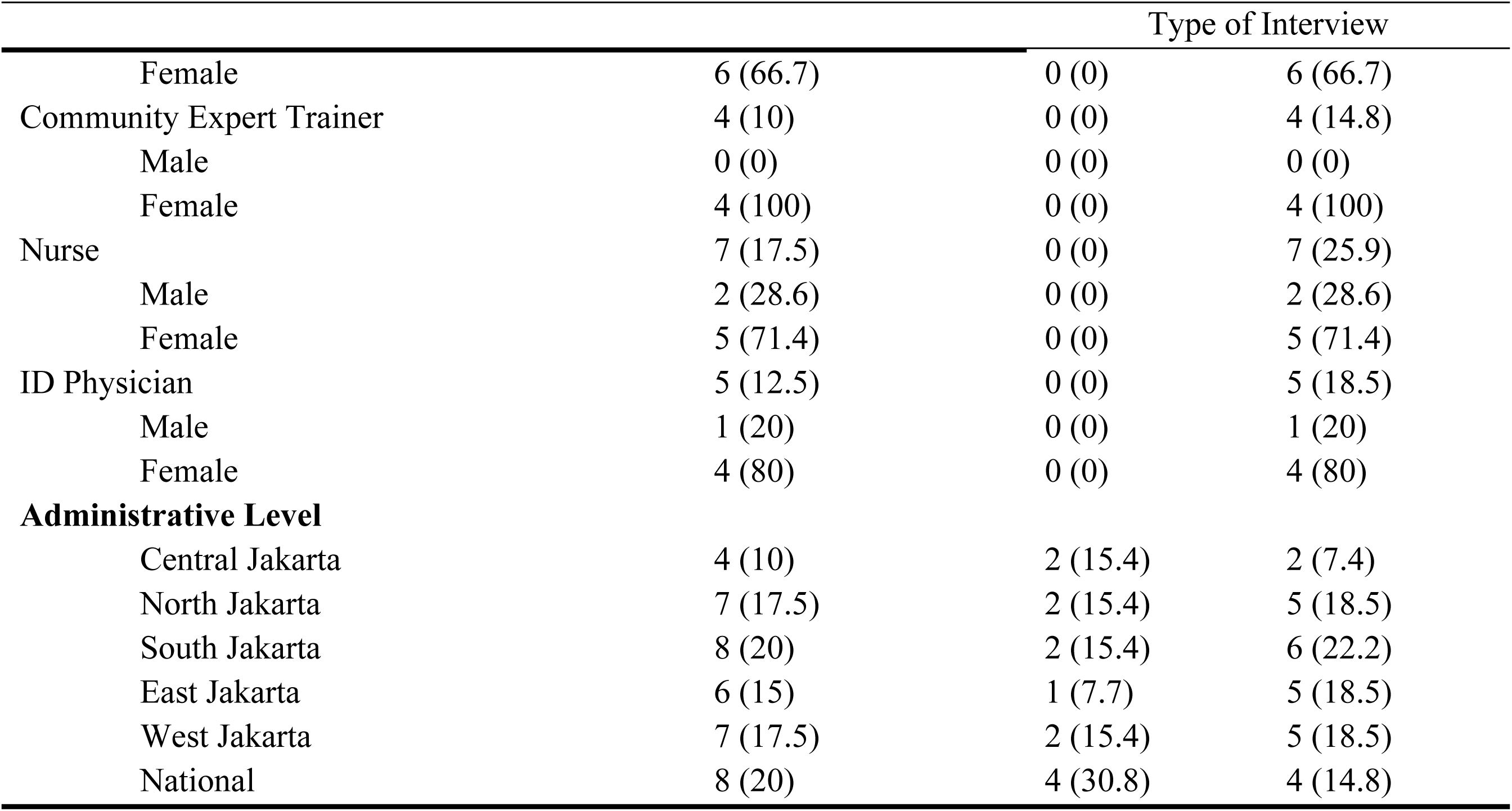
Characteristics of IDI and FGD Participants.

The interview participants were categorised by gender, diversity, organisation, position, and administrative level. Males constituted 20% of the sample, with higher representation in IDIs (38.5%) than in FGDs (11.1%). Females comprised 80%, represented in IDI (61.5%) and FGD (88.9%). Policymakers comprised 17.5%, international partners 5%, and CETs 10% of participants. Healthcare workers represented 67.5%, with a similar distribution in the IDI (69.2%) and FGD (66.7%). Most participants (85%) were affiliated with the Indonesian MoH. HIV Programme Coordinators constituted 32.5% of the participants, predominantly in FGDs (40.7%) versus IDIs (15.4%), with 92.3% being female. Heads of Primary Health Centres (22.5%) represented 69.2% of the IDI participants. Nurses (17.5%) and ID physicians (12.5%) comprised 44.4% of the FGD participants.

### CBPA in Planning, Adapting and Implementation

The IPSD intervention involved forming a diverse module writing team, including community representatives, healthcare providers, Health Department officials, UN agencies, and the Development and Empowerment of Health Human Resources agency. *“The module writing team consisted of community members, healthcare providers as trainers, Provincial Health Office representatives, and our team members. A Provincial Health Office representative communicated with the Ministry of Health’s Development and Empowerment of Human Resources agency.”* (Male, Project Director, FHI360).

All subgroups of PHIV-KPs were represented in the module development process. This multi-stakeholder collaboration ensured that the IPSD training module reflected a comprehensive understanding of how stigma and discrimination are manifested and experienced by different KP communities in Indonesia. The multi-stakeholder collaborative approach was confirmed by a CET:

> Back then, those who provided input for the module were [name of transgender], [name of MSM-1], the late [name of MSM-2], and if I am not mistaken, there were seven people… The collaboration was very good because they (MoH) managed to gather a significant number of hospitals to actively participate in training… it was really cool to see them participate. (Female, Community Expert Trainer, FGD 2)

The IPSD module underwent limited trial testing with community representatives, allowing for feedback and adjustments based on real experiences. The flexibility of the intervention enabled content optimisation for learning outcomes. *“During pilot training, we simplified complex translated sentences while maintaining the original meaning.”* (Female, Provincial-level Policymaker).

Module adaptation was conducted by multidisciplinary experts and during pilot testing. Trainers were selected from diverse backgrounds, including KP experts, healthcare workers, and social service representatives. CETs handled stigma, discrimination, and SOGIE modules, whereas policymakers and healthcare professionals covered epidemiology, clinical practice, and government policies.

> We covered HIV, STIs, stigma, discrimination, and SOGIE topics. *Representatives from the Health Department, City Health Office, and EPIC were present as trainers. When the community delivers messages in the presence of government agencies, it strengthens their intervention.* As policymakers, our presence makes policies and information more trusted with government agencies and community representatives. (Female, Provincial-level Policymaker)

This statement highlights the importance of government-led initiatives in disseminating interventions and addressing sensitive issues affecting HIV treatment in healthcare facilities. The presence of policymakers and physicians adds credibility to the delivery of IPSD training modules. A multidisciplinary training team enables discussions on HIV regulations and protocols, enhancing information credibility.

CBPA facilitates IPSD implementation through community perspectives and patient-centred care practices. *“The response exceeded expectations; health office participants appreciated the model. The community trainers connected discussions with real experiences, making sessions livelier.”* (Male, Project Director, FHI360).

HCWs were more receptive to receiving information delivered by community trainers, thus creating a more trusted and relatable learning environment (Table 2).

**Table 2:**
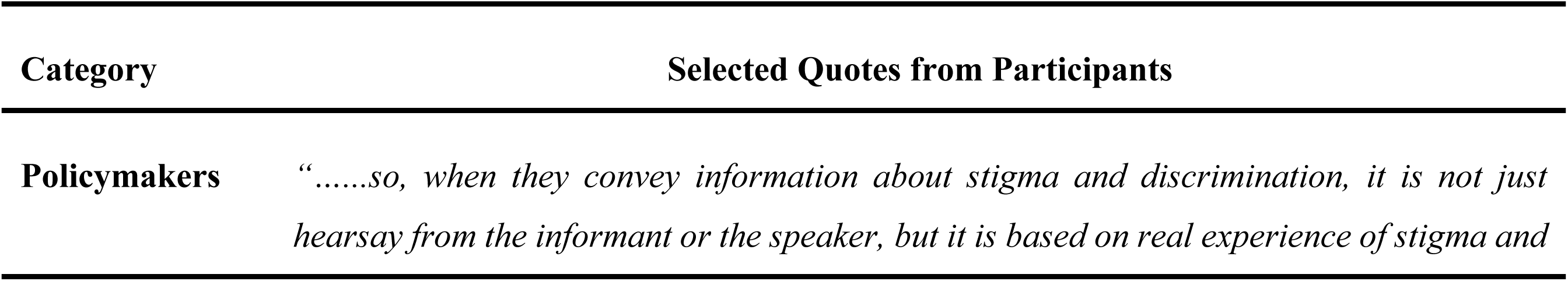
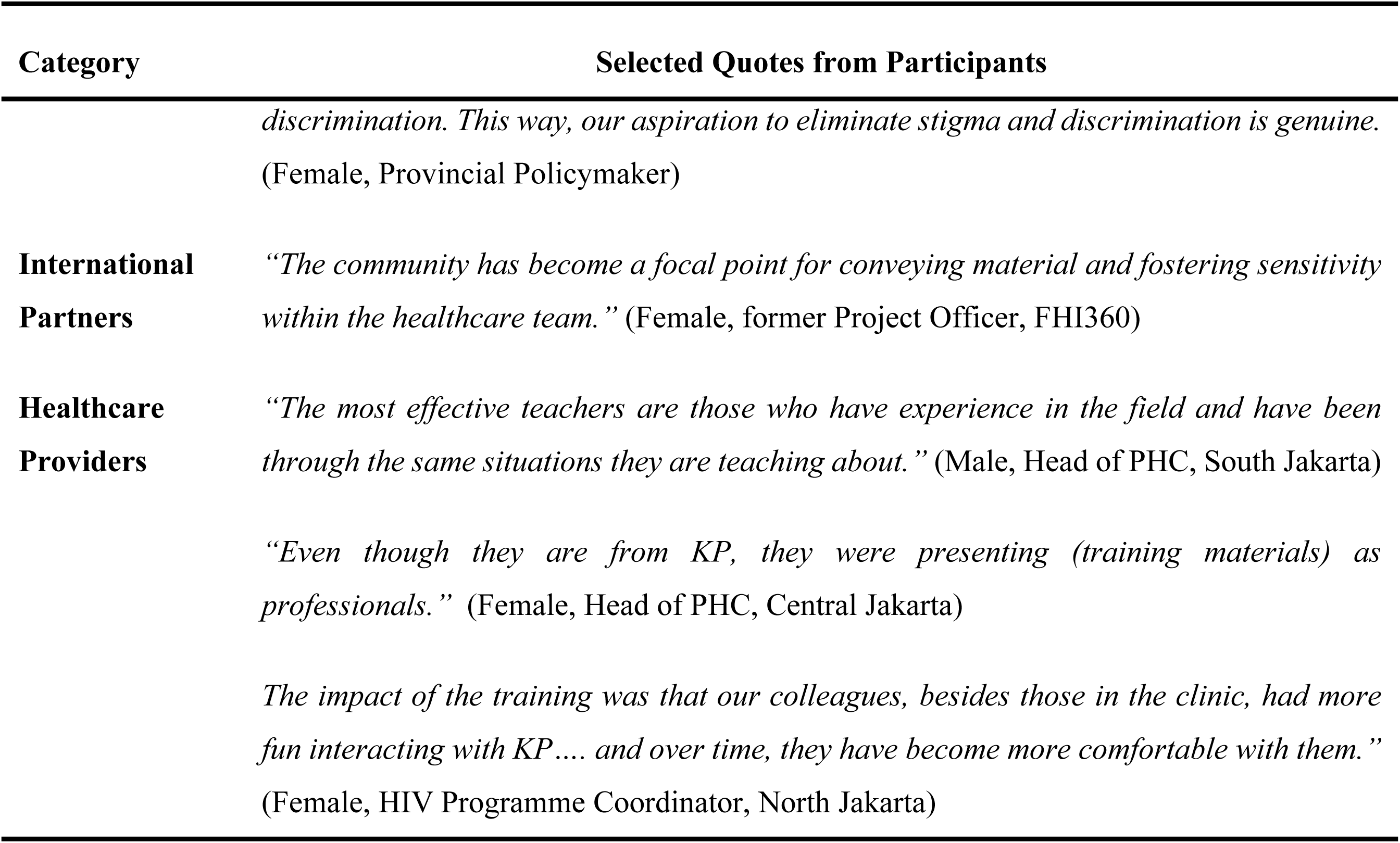
Quotes to Illustrate Participants’ Response to CET.

| Category | Selected Quotes from Participants |
| --- | --- |
| Policymakers | *".....so, when they convey information about stigma and discrimination, it is not just hearsay from the informant or the speaker, but it is based on real experience of stigma and* |
|  | *discrimination. This way, our aspiration to eliminate stigma and discrimination is genuine.*<br>(Female, Provincial Policymaker) |
| **International Partners** | *"The community has become a focal point for conveying material and fostering sensitivity within the healthcare team."* (Female, former Project Officer, FHI360) |
| **Healthcare Providers** | *"The most effective teachers are those who have experience in the field and have been through the same situations they are teaching about."* (Male, Head of PHC, South Jakarta) |
|  | *"Even though they are from KP, they were presenting (training materials) as professionals."* (Female, Head of PHC, Central Jakarta) |
|  | *The impact of the training was that our colleagues, besides those in the clinic, had more fun interacting with KP.... and over time, they have become more comfortable with them."*<br>(Female, HIV Programme Coordinator, North Jakarta) |

#### Breaking Down Barriers and Dispelling Fears

CETs are living examples of KP who thrive with HIV, demonstrating they can lead good lives while adhering to treatment and maintaining undetectable viral loads, “*People with personal experiences become powerful real-life examples that impact others’ thinking.”* (Female, Head of PHC, Central Jakarta, IDI 20)

By maintaining treatment adherence and leading fulfilling lives, CETs challenge stereotypes associated with PHIVs. Sharing positive encounters with healthcare providers helps correct negative perceptions of both service providers and recipients. Respondents noted initial awkwardness with CETs, showing preexisting stereotypes: *“some of my colleagues had experiences like before meeting them (CETs), they were like, “How will it be?” Some felt tense or awkward. But after meeting them, there was nothing to be afraid of.”* (Female, HIV Coordinator, West Jakarta).

Personal interactions with CETs effectively desensitised HCWs and created empathy. As one participant noted, *“it turns out that it (IPSD) changes the mindset of the health centre staff… with the participation of KPs, the barriers to the adoption of IPSD seem to disappear.”* (Female, Head PHC, West Jakarta).

Language sensitivity is crucial, particularly regarding the preferred names and terms used by transgender people and MSM.

> When I attended the training, I took notes on specific terms used by transgender people and MSM. It is essential to be aware of phrases like “Oh dendong (to dress up), oh cucok, ditempong (anal sex)” so that we know how to communicate effectively. If the healthcare providers don’t understand such language, they might unintentionally reject them. (Female, Head of PHC, West Jakarta)

IPSD training allowed participants to explore issues beyond stigma, highlighting the importance of understanding the impact of multiple levels of stigma on health outcomes and comprehensive care models, including mental health support for key populations.

CBPA, as an implementation strategy, promotes the principles of equality and the recognition that there is inherent value in everyone, worthy of respect and importance. The following statement strongly advocates the CBPA strategy.

> The community has played a significant role in transforming healthcare workers’ attitudes and perceptions. *Through community involvement, healthcare workers who previously exhibited phobia or discomfort towards KP individuals become aware of and acknowledge the presence of sexual diversity in society.* (Female, former Project Officer, FHI360)

#### Authenticity of Real-Life Examples

One key feature of IPSD training is the use of Expert Version Testimony (EVT) as an effective tool to communicate first-hand experiences of stigma and discrimination encountered by KPs. CETs’ sharing sessions had a profound impact on influencing HCW’s perspectives by helping participants connect at a personal level with the challenges faced by PHIV. Storytelling and real-life examples were highlighted by participants as particularly impactful (Table 3).

**Table 3:** Quotes to Illustrate Authenticity of Real-Life Examples.

| Category | Selected Quotes from Participants |
| --- | --- |
| **Policymakers** | *"In other regions as well, the acceptance is good when facilitators come from NGOs, as they can provide direct testimonials based on their own experiences as individuals living with HIV."* (Female, HIV Programme Coordinator, South Jakarta)<br>*"In my opinion, it serves as a supportive factor because individuals from KP provide testimonials like, "I am from the KP, I have been living with HIV, and I am doing fine." So, the main point is to ensure that service providers and the community can accept that it's safe to interact with PHIV. They can learn from these experiences and reflect upon them during (when providing) HIV testing."* (Female, HIV Programme Coordinator, East Jakarta) |
| **Healthcare Providers** | *"If the community members are truly immersed in the situation, then I think we can actually gain more value from them because they are directly involved. Surely, there are values that we may not obtain from colleagues. Finally, we obtain it from community members."* (Female, ID Physician, East Jakarta)<br>*"The idea is that the message being conveyed might be better delivered by community members who belong to a KP. For example, in the case of transgender individuals, understanding their fears and expectations regarding healthcare services wouldn't be an issue if the material is presented by members of their community."* (Female, HIV Programme Coordinator, South Jakarta) |
|  | *We incorporate what we know as healthcare workers by putting ourselves in their shoes, by being in their position, and pretending to be a part of them so that we can feel and empathize with them. (Male, Head of PHC, North Jakarta)* |
|  | *" The most effective teachers are those who have fieldwork experience and personal knowledge of the materials they are teaching about. This was very suitable because they could share their stories of how they overcame their challenges. They would say things like, "I used to be in such a bad state, and it was really hard to get out of it, but with determination, commitment, and the goal of seeking treatment, I managed to turn my life around over time." (Male, Head of PHC, South Jakarta)* |

The following statement compares training by doctors who provide theoretical knowledge with learning from CETs with lived experiences of stigma.

> So, having these speakers who had real-life experiences and had successfully gone through those challenges was impactful. It was better than the theoretical knowledge of doctors. When training comes from doctors, it is often just listening, and we might not fully grasp it, tending to protest when implementing it. *We may teach it but not practice it.* (Male, Head of PHC, South Jakarta)

The participant believes that information delivery about HIV stigma by healthcare professionals is less effective than that by CETs who have experienced stigma themselves. CETs are more effective in delivering stigma, discrimination, and SOGIE modules, facilitating behavioural change among HCWs, and creating a more supportive environment for PHIV and key populations.

Healthcare workers view CETs as positive transformative external change agents who can influence their peers’ health-seeking behaviours. Outside the clinical setting, CETs are acknowledged as community leaders and long-standing HIV advocates.

## Discussion

This study identified a gender disparity among the participant groups. In FGDs, 89% of participants were women, highlighting the predominance of females in Indonesian healthcare, particularly in patient-care roles. Conversely, IDIs featured a higher proportion of men, suggesting greater representation in leadership positions among policymakers and funding agencies. Women constituted 61.5% of the IDI participants and 88.9% of the FGDs. Among the healthcare workers, 80% were women. Data from the Ministry of Health corroborate this trend, indicating that women comprise 70.9% of Indonesia’s health workforce, with complete dominance in midwifery (100%) and a significant presence in Healthcare Services (80%). However, men are more prevalent in Disease Prevention (57.8%) and medical specialties (60.8%), underscoring their dominance in high-level positions and specialised fields (22).

This study demonstrates how the CBPA can significantly enhance the adoption of health interventions in low-resource settings. This provides evidence that sustainable change in stigma reduction is not only possible but inevitable when interventions are co-created with affected communities and executed through trusted community networks.

External Change Agents, particularly Community Expert Trainers (CETs), emerge as pivotal figures in this process, acting as influential opinion leaders who dismantle entrenched stereotypes. The dynamic collaboration between the PHIV and KP communities as External Change Agents, alongside FHI 360, has revolutionised the IPSD training curriculum and its implementation. This strategic alliance has ensured the module’s relevance and alignment with community insights and adhered to government-endorsed practices, making it a model of excellence. Through the expertise of PHIV and KP, the CBPA initiative has profoundly deepened our understanding of lived experiences, fostering critical discourse on SOGIE, stigma, and discrimination. The real-life narratives shared by CETs cultivated a trusted learning environment for healthcare workers, significantly diminishing their phobias and discomfort.

The active participation of PHIV, KP, healthcare providers, and stakeholders in crafting this intervention underscores the unparalleled effectiveness of collaborative public health approaches. The CBPA strategy for the IPSD intervention is a beacon of progress, promoting improved attitudes among non-stigmatised individuals by facilitating meaningful interactions with the stigmatised groups (23). It champions implementation designs that enhance interactions between individuals with stigmatised conditions and stakeholders (3) and unequivocally demonstrates how educational efforts with structured contact programs yield the most impactful results (24).

### Policy Implications

In Indonesia, the findings present a compelling case for the government to decisively allocate public health resources and make policy-level decisions prioritising community-led responses. This is especially crucial for interventions addressing human rights and gender intersectionality in HIV programs. Recognising the indispensable role of community-led responses through CBPA not only underscores the community’s significance as pivotal stakeholders in the national HIV response but also highlights their unparalleled capacity and expertise in effectively driving stigma reduction initiatives.

By demonstrating how the CBPA has successfully facilitated the adoption of interventions at provincial and district-level health facilities, PWH and KP communities are empowered to vigorously advocate for the government and international agencies to prioritise community participation in tackling the critical findings from the Stigma Index 2.0 study.

### Strengths and Limitations

The study design enhances the validity and supports the hypothesis. The CFIR domains shaped the semi-structured interview questions, enabling deductive analysis with CFIR constructs as the theoretical framework. Through iterative thematic analysis, patterns corresponding to the CFIR constructs were identified (25). This study examined the IPSD implementation context, considering barriers and facilitators affecting outcomes. Semi-structured interviews allowed the exploration of new directions based on participant feedback, uncovering issues such as community expert trainer motivations. This qualitative approach captured the implementers’ experiences in maintaining the intervention.

This study faced recruitment challenges. An IDI with a PHC Head was impossible due to scheduling conflicts, and a planned FGD with district-level KP experts could not proceed as many reduced their involvement after transitioning online during COVID-19. However, the FGD with KP experts included participants active in national and district-level health authorities, although limited representation may have affected the understanding of certain contextual aspects.

## Conclusion

This study presents evidence supporting the Community-based Participatory Approach (CBPA) as an implementation strategy and highlights the essential role of External Change Agents in advancing CBPA as an effective strategy for implementing HIV stigma reduction interventions in healthcare settings. These findings have substantial policy implications, strongly advocating for the integration of community participatory approaches into HIV programming. By incorporating the invaluable perspectives of PWH and KPs, the CBPA strategy ensures that the IPSD intervention is not only relevant but also contextually tailored to meet the needs of those it serves.

This research lays the foundation for future exploration of stigma reduction and implementation strategies, making a compelling case for investing in capacity building to propel implementation science in Southeast Asia. By doing so, healthcare systems will be empowered to deliver impactful interventions that decisively address stigma in HIV care.

## Data Availability

The datasets generated and/or analysed during the current study are not publicly available due to confidentiality agreements and ethical restrictions involving participant privacy. However, de-identified data may be made available from the corresponding author upon reasonable request and with approval from the relevant ethics committee. Any other identifying information related to the authors and/or their institutions, funders, approval committees, etc, that might compromise anonymity.

## Supporting Information

S1 Appendix 1: Semi-structured Interview Guide in English and Bahasa Indonesia

S2 Appendix 2: Informed consent

**Figure I:**
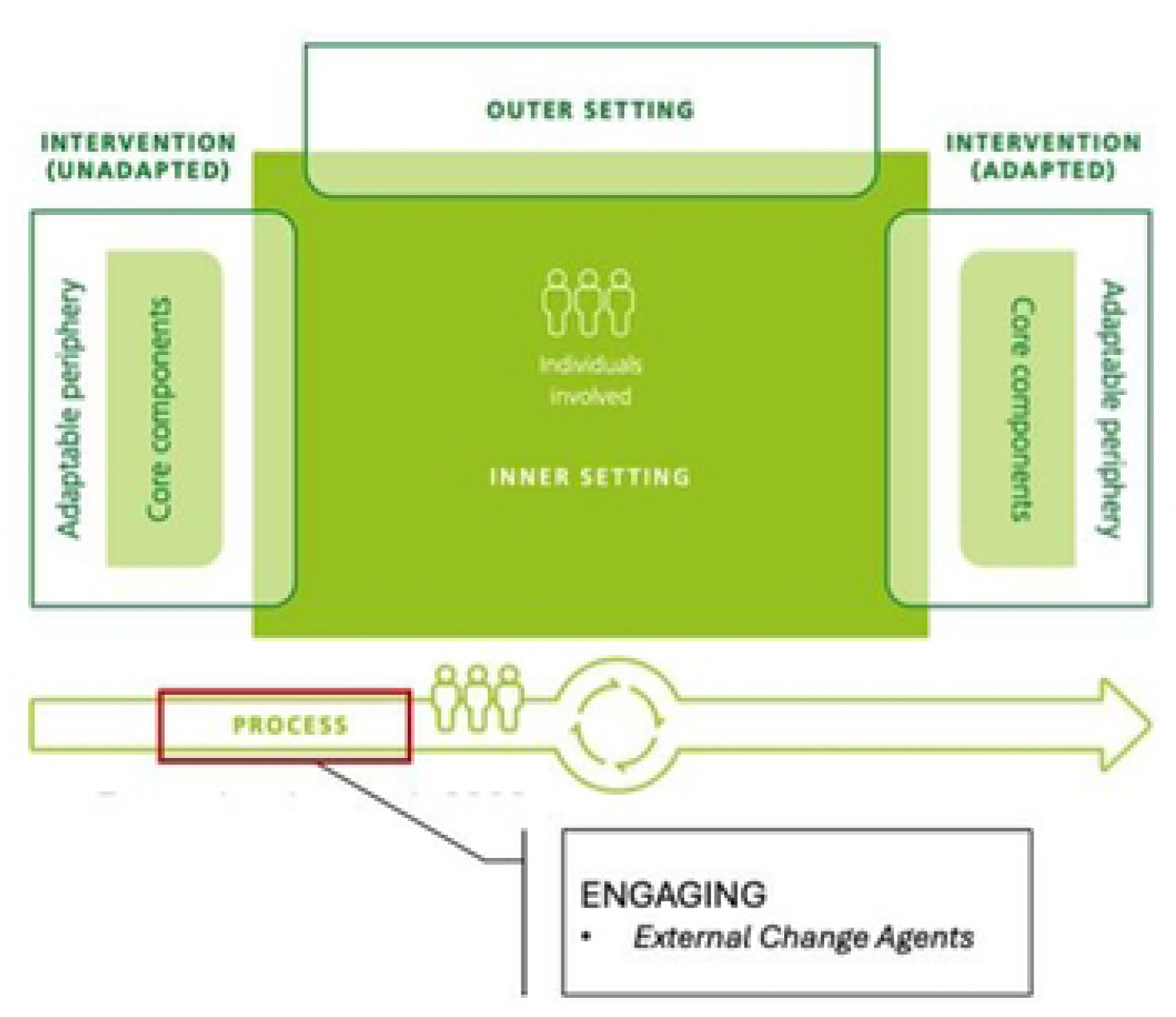
CFIR Framework-Process (Domain), Engagine (Construct). External Change Agent (Sub-construct)

